# Fractional tumor burden refines prognostic stratification beyond MGMT status in glioblastoma after chemoradiotherapy

**DOI:** 10.64898/2026.10.29.26366540

**Authors:** Melissa A. Prah, Jennifer M. Connelly, Kathleen M. Schmainda

**Affiliations:** Department of Biophysics, Medical College of Wisconsin, Milwaukee, WI, United States; Department of Neurology, Medical College of Wisconsin, Milwaukee, WI, United States; Department of Radiology, Medical College of Wisconsin, Milwaukee, WI, United States

## Abstract

**Background and Purpose:** Following standard-of-care treatment for glioblastoma, response to chemoradiotherapy is difficult to evaluate because changes on postcontrast T1WI may represent either treatment effect or tumor progression, particularly in the early posttreatment period. Although O6-methylguanine DNA methyltransferase promoter methylation is prognostic and informs treatment response, outcomes among patients remain heterogeneous. We hypothesized that combining advanced physiologic imaging with methylation status would improve early prognostic stratification.

**Materials and Methods.:** In this retrospective cohort study, patients with newly diagnosed glioblastoma who underwent surgical resection or biopsy followed by standard chemoradiotherapy and had diffusion- and perfusion-weighted MR imaging within 6 weeks after treatment completion were included. Imaging-derived metrics included mean apparent diffusion coefficient (ADC), mean standardized relative cerebral blood volume (sRCBV), change in contrast-enhancing volume on dT1, and the volume of fractional tumor burden (FTB), which is the volume of enhancing tissue with moderate-to-high probability of vascular tumor. Overall survival was evaluated using Kaplan-Meier analysis with log-rank testing.

**Results:** Forty-three patients (21 male, 22 female) were included with a mean age of 57 years. Median overall survival was longer in patients with O6-methylguanine DNA methyltransferase-methylated tumors compared with unmethylated tumors (19.7 vs 10.6 months; P = .01). Stratification by mean sRCBV, mean ADC, or change in dT1 volumes was not associated with overall survival (P > .05 for all comparisons). Among unmethylated tumors, FTB <5 mL was associated with longer overall survival compared with ≥5 mL (14.5 vs 8.4 months; P = .006), whereas no survival difference, based on FTB volumes, was observed in methylated tumors (P = .50).

**Conclusions:** Fractional tumor burden, measured within 6 weeks after chemoradiotherapy, refines prognostic stratification beyond methylation status in glioblastoma, identifying a subgroup of unmethylated tumors with longer overall survival. These findings support the use of spatially resolved perfusion imaging to improve interpretation of early posttreatment enhancement.

## INTRODUCTION

Survival for patients with a newly diagnosed high grade glioma remains dismal, with significantly worse outcomes reported for those who possess the isocitrate dehydrogenase (IDH1/2)-wild-type genetic variant.^1,2^ For these more aggressive World Health Organization (WHO) grade 4 tumors, current standard of care includes maximal safe resection followed by radiation therapy with concomitant chemotherapy (CRT) followed by adjuvant chemotherapy along with optional tumor treating fields.^3^ While age at diagnosis, extent of resection, and receipt of CRT may influence overall survival (OS), one of the most prognostically relevant biomarkers for patients treated with standard of care is methylation of the O6-methylguanine DNA methyltransferase (MGMT) promoter, where those with MGMT-methylated tumors demonstrate significantly improved OS compared with those with an unmethylated promoter^4–6^. In patients with an unmethylated MGMT promoter, the DNA repair enzyme is expressed and therefore promotes resistance to the cytotoxic effects of the alkylating chemotherapeutic agent temozolomide.^7^ Beyond chemotherapy, MGMT status has also been reported to impact patients receiving irradiation alone, where a twofold increase in progression during radiation treatment among tumors with an unmethylated MGMT promoter has been reported.^8^

While MGMT promoter methylation testing is an invaluable prognostic tool for identifying the roughly 40% of patients with MGMT-methylated tumors, who are more likely to benefit from CRT, the therapeutic course remains the same for both methylated and unmethylated MGMT populations; the exception being that the European Association of Neuro-Oncology reviews MGMT status when considering temozolomide use in elderly patients.^9,10^ Therefore, outside of clinical trial design and drug development efforts to alter MGMT function, MGMT methylation status is primarily utilized as a clinical tool alongside imaging for improved confidence in the evaluation of response or progression after treatment with CRT.^9^

Following CRT, upwards of 20% of tumors display a radiographic increase in contrast enhancement on MR T1WI, termed pseudoprogression, thought to represent a positive biological response.^11,12^ This radiographic change typically occurs within three months of CRT completion and has been largely associated with MGMT methylated GBM, observed to a minor extent in MGMT unmethylated GBM, and shown to be predictive of prolonged OS when present.^13,14^

While T1WI cannot effectively distinguish early post-CRT response from progression, advanced perfusion imaging such as DSC MR imaging evaluates tumor vascularity and therefore is not subject to the same false appearance of progression seen on conventional contrast-enhanced imaging. DSC-derived relative CBV serves as a surrogate marker of tumor vascularity and has been shown to correlate with glioma grade, tumor aggressiveness, and treatment response.^15–21^ Building on this, fractional tumor burden (FTB)^12,21–23^ applies voxel-wise classification of relative CBV values to quantify the proportion of enhancing tissue consistent with viable tumor. FTB has been shown to be clinically useful for both visual and quantitative assessment of tumor progression and response.^12,21,23–25^

Additionally, advanced MR imaging such as DWI-ADC has potential for identification of post-CRT response from tumor progression, where higher ADC values have been reported in those exhibiting pseudoprogression on T1WI.^26^ However, as ADC is inversely correlated to tumor cellularity, it may be more prone to false negatives in regions of hypercellularity mixed with edema and inflammation.^27^ While noting that DSC may hold greater clinical potential due to its technical feasibility, availability, and short acquisition time, a recent review of published studies identified both DWI and DSC as highly accurate for discriminating T1WI pseudoprogressive response from progression, with an area under the curve of 0.93 for both methods.^28^

Because MGMT methylation status reflects underlying tumor biology while advanced MR imaging biomarkers provide physiologic measures of tumor characteristics such as vascularity and cellularity, integration of molecular and imaging biomarkers may improve prognostic stratification following treatment. While MGMT methylation is associated with improved OS, outcomes among patients with unmethylated MGMT remain heterogeneous and are not well stratified by existing biomarkers.^9^ Accordingly, the relationship between MGMT methylation status and advanced MR imaging biomarkers such as DSC-derived fractional tumor burden has not been well characterized. Therefore, the purpose of this study was to determine whether advanced MR imaging biomarkers, including DSC-derived fractional tumor burden and ADC, provide additional prognostic stratification beyond MGMT methylation status with respect to OS following chemoradiotherapy.

## METHODS

### Participants

This retrospective observational cohort study was conducted in accordance with the STROBE reporting guidelines. Between 2009 and 2019, participants were consecutively identified from a perfusion imaging database. All patients meeting the study inclusion criteria were considered for analysis. Written, informed consent was obtained for all participants following institutional review board approval in accordance with HIPAA guidelines. The number of included participants was based on available data and not upon a predetermined power analysis. To be considered for inclusion, patients were required to have undergone surgical resection or biopsy of a newly diagnosed GBM and received standard postoperative treatment including CRT and adjuvant temozolomide with or without tumor treating fields. Participants were required to have advanced imaging including DWI and DSC MR imaging within six weeks after completion of CRT, where only technically adequate imaging datasets permitting generation of the required parametric maps were included.^29^ Histopathologic diagnoses were established clinically according to current WHO classification at the time of diagnosis. Patients with unknown MGMT methylation status and known IDH1/2-mutant glioma were not considered for inclusion. MGMT methylation was determined as part of the clinical record. Additionally, only patients with measurable disease comprising at least 1 mL of residual contrast enhancement on postcontrast T1WI following CRT completion were included.

### Imaging acquisition and processing

All MR imaging examinations were performed on 1.5T or 3T systems. DSC perfusion imaging was acquired using a gradient-echo EPI sequence following a preload dose of gadolinium-based contrast agent, with the bolus administered 60 seconds after the start of image acquisition according to the prescribed protocol^29,30^: TE = 30 ms, TR = 1,250 ms, flip angle = 60°, slice thickness = 5 mm, interslice gap = 1.5 mm, matrix = 96-128 x 96-128, FOV = 220-240 x 220-240 mm^2^. Isotropic DWI was acquired with the following prescribed settings: b-values = 0 and 1000 s/mm^2^, TE = 80 ms, TR = 8,000 ms, flip angle = 90°, slice thickness = 3 mm, interslice gap = 1 mm, matrix = 128 x 128, FOV = 110 x 110 mm^2^.

Maps of dT1, DWI-derived ADC, and leakage-corrected DSC-derived standardized rCBV (sRCBV) were generated using OsiriX MD (Pixmeo, Bernex, Switzerland) and the Imaging Biometrics LLC (Elm Grove, WI) suite of FDA-cleared plug-ins.^15,29–32^ For dT1 map generation, precontrast T1WI was first co-registered to postcontrast T1WI. Standardized precontrast T1WI were then subtracted from standardized postcontrast T1WI to generate dT1 maps.^33^ This approach minimizes variability related to acquisition parameter settings and field strength, provided that precontrast and postcontrast T1WI are acquired with matched sequence parameters within each exam. This method also suppresses signal contributions from bright pre-contrast signal on T1WI, due to blood products and proteinaceous material, while preserving contrast enhancement associated with tumor.

Post-CRT contrast-enhancing lesion volumes were delineated and quantified by applying a predefined threshold of 3,000 to the dT1 map within a manually defined bounding region that encompasses the lesion. The same threshold was applied uniformly across all subjects. The dT1 volumes enable assessment of changes in the enhancing volume following chemoradiotherapy, reflecting the standard approach of determining treatment response. In addition to the contrast-enhancing lesion, a region of interest encompassing the hyperintense lesion on FLAIR imaging was manually defined to capture the broader region of tumor-related abnormality.

Maps of sRCBV were generated using intensity standardization available within the Imaging Biometrics platform (Imaging Biometrics LLC, Elm Grove, WI). By eliminating the need for a normalizing ROI and reducing dependence on slight variations in acquisition parameters, field strength, and vendor platforms, for example, sRCBV has demonstrated improved repeatability across patients and between exams.^32^ All parametric maps were subsequently co-registered to postcontrast T1WI. Mean sRCBV values were extracted from the dT1 ROI, whereas mean ADC values were extracted from the FLAIR-defined lesion ROI.

FTB maps were generated using sRCBV thresholds.^23^ Specifically, sRCBV values within the dT1-defined contrast-enhancing region were thresholded at levels of 1.00 and 1.556, corresponding to low (sRCBV <1.00), moderate (sRCBV: 1.00-1.556), and high (sRCBV >1.556) probability of tumor.^22^ These regions are colorized as blue, yellow and red on FTB maps (Figure 1). FTB volume was defined as the combined volume of voxels classified as moderate- and high-probability of tumor (yellow + red). Figure 1 shows representative multiparametric imaging and derived parameter maps evaluated in this study.

**Figure 1.**
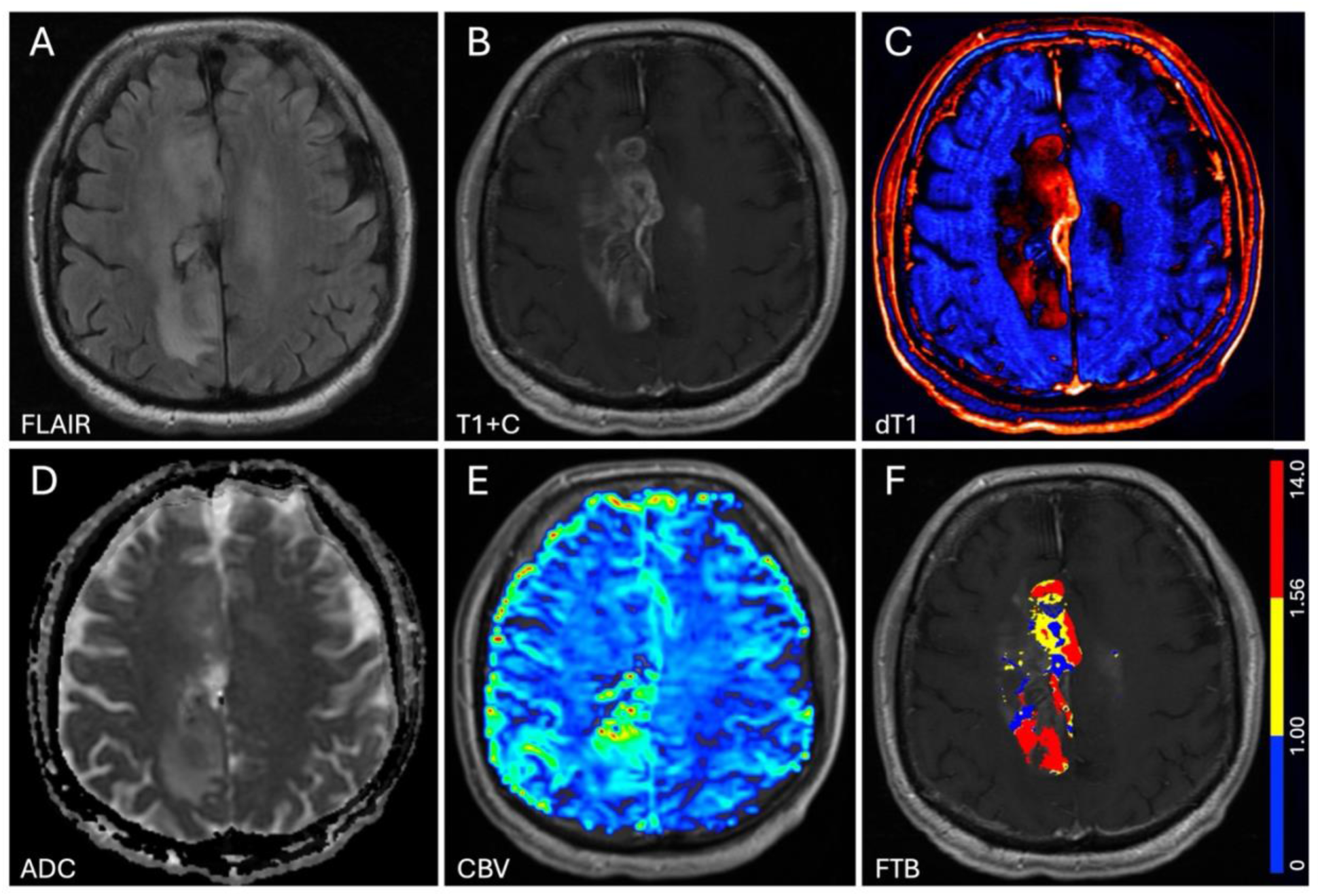
Representative multiparametric MR imaging demonstrating spatial correspondence between conventional MRI, including (A) FLAIR and (B) postcontrast T1WI, and derived maps including (C) ΔT1, (D) ADC, (E) standardized relative CBV, and (F) fractional tumor burden (FTB). FTB is calculated within the enhancing ΔT1 lesion and demonstrates the spatial distribution of low (blue), moderate (yellow), and high (red) tumor probability.

### Statistical Analysis

OS was calculated from the post-CRT imaging timepoint to death, or last follow-up, and evaluated over a 24-month follow-up period. Survival distributions were estimated using the Kaplan-Meier method and compared using the log-rank test.

Stratification of sRCBV and ADC was based on the population median. Change in dT1 contrast-enhancing lesion volume from pre-CRT baseline to the first post-CRT imaging timepoint was evaluated as a marker of treatment response by dichotomizing patients according to negative versus positive change relative to baseline lesion volume. Fractional tumor burden was stratified empirically using a threshold of 5 mL.

All analyses were performed using GraphPad Prism (GraphPad Software, Boston, Massachusetts). A 2-sided P value < .05 was considered statistically significant.

## RESULTS

Data from 43 participants (21 male, 22 female) with a mean age of 57 (23-74) years were available for analysis. MR imaging was acquired at a mean of 29 (12-42) days following completion of CRT. Surgical management consisted of tumor resection in 42 participants and biopsy in 1 participant. Ten patients also received tumor treating fields as part of adjuvant therapy. During the study period, 33 deaths were observed. Across the full cohort, imaging-derived metrics demonstrated a median ADC within the FLAIR-defined lesion ROI of 1,331.1 ×10^-^⁶ (502.0-1,552.6 ×10^-^⁶) mm²/s and a median sRCBV within the dT1 contrast-enhancing lesion ROI of 1.34 (0.43-3.24). The mean dT1 contrast-enhancing lesion volume was 15.28 (1.34-72.73) mL, and the mean fractional tumor burden volume was 9.61 (0.08-70.48) mL.

Kaplan-Meier analysis demonstrated significantly longer OS in patients with MGMT-methylated tumors (n = 17) compared with those with unmethylated tumors (n = 26) (median OS, 19.67 vs 10.59 months; log-rank P = .01) (Figure 2).

**Figure 2.**
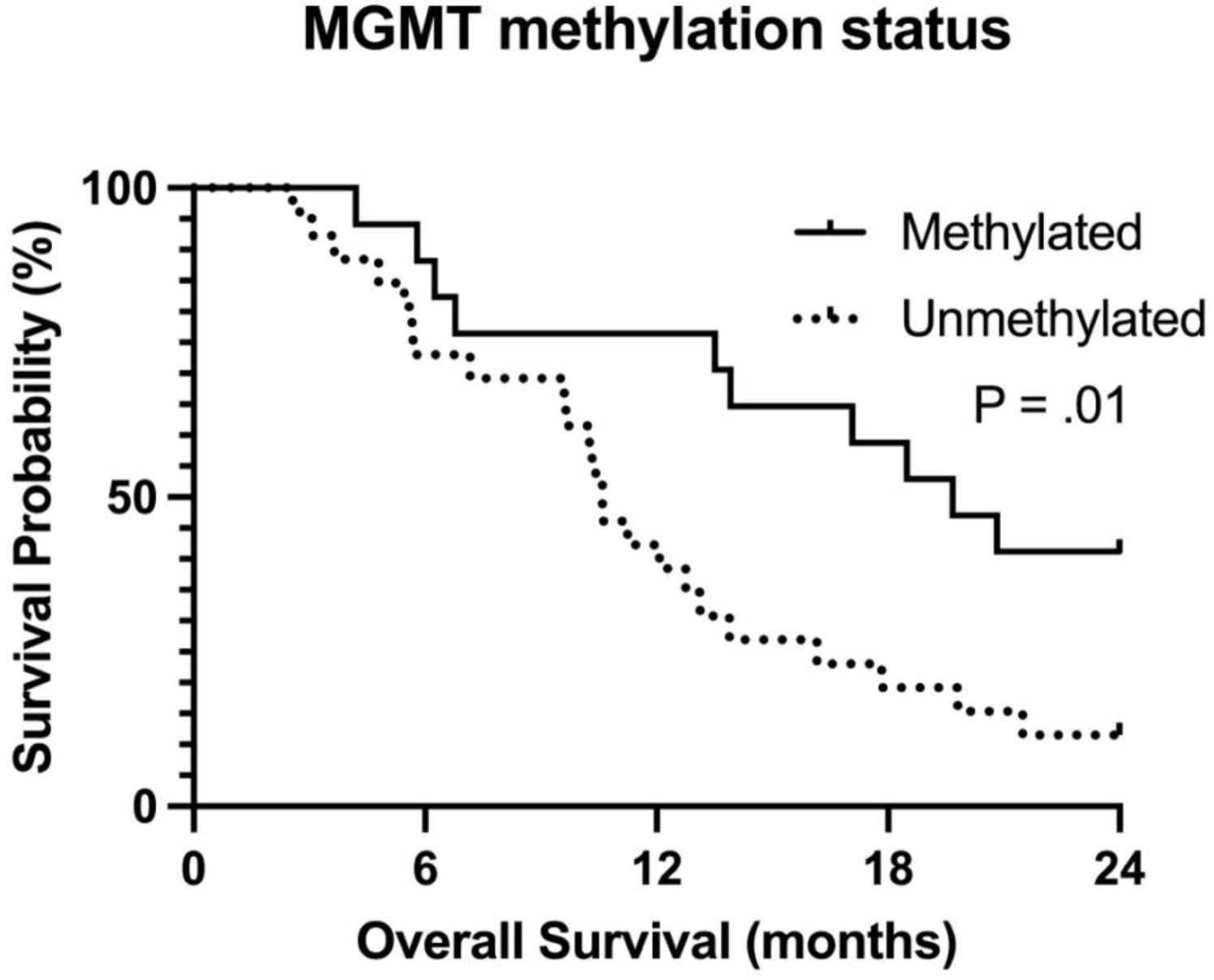
Kaplan-Meier analysis of OS stratified by MGMT methylation status. Patients with MGMT-methylated tumors demonstrated significantly longer OS compared with those with MGMT-unmethylated tumors (median OS, 19.67 vs 10.59 months; log-rank P = .01).

Stratification by sRCBV or ADC using the population median did not demonstrate significant differences in OS for either MGMT-unmethylated or methylated tumors (sRCBV: P = .52 and P = .90; ADC: P = .25 and P = .23, respectively). One participant was excluded from the dT1 percent change analysis due to the absence of a baseline pre-CRT T1WI exam.

Similarly, a binary change in dT1 contrast-enhancing lesion volume from baseline to the first post-CRT imaging timepoint was not associated with OS in either MGMT-unmethylated or methylated tumors (P = .17 and P = .14, respectively).

In contrast, among MGMT-unmethylated tumors, OS was significantly longer when FTB volume was <5 mL compared with ≥5 mL (median OS, 14.46 vs 8.39 months; log-rank P = .006) (Figure 3A). However, for methylated tumors, no significant differences in OS for FTB volume stratification were observed (median OS, 18.49 vs 21.84 months; log-rank P = .50) (Figure 3B).

**Figure 3.**
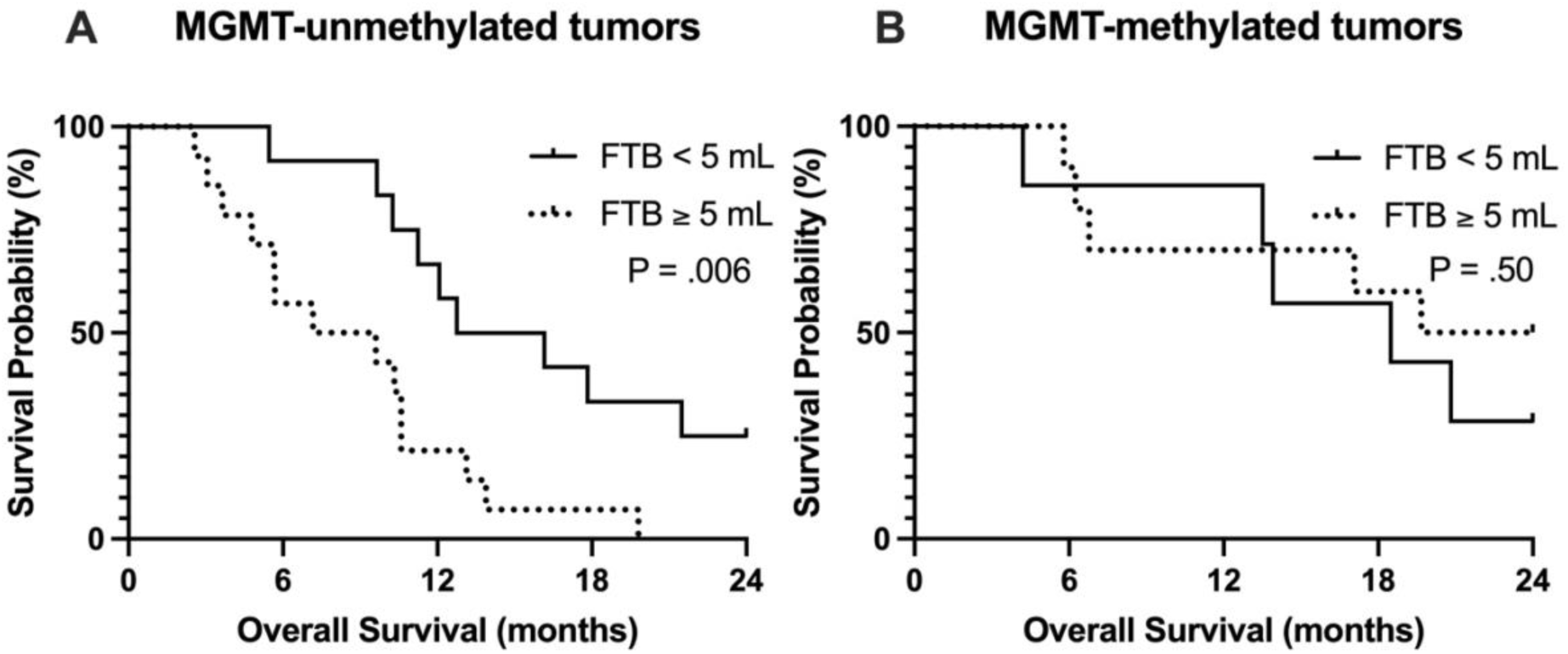
Kaplan-Meier analysis of OS stratified by fractional tumor burden (FTB) volume in (A) MGMT-unmethylated and (B) MGMT-methylated tumors. Lower FTB volume (<5 mL) was associated with improved OS in MGMT-unmethylated tumors (median OS, 14.46 vs 8.39 months; log-rank P = .006), whereas no significant difference in OS was observed in MGMT-methylated tumors (median OS, 18.49 vs 21.84 months; log-rank P = .50).

Representative FTB maps are shown in Figure 4, illustrating FTB volume relative to OS in both methylated and unmethylated subjects.

**Figure 4.**
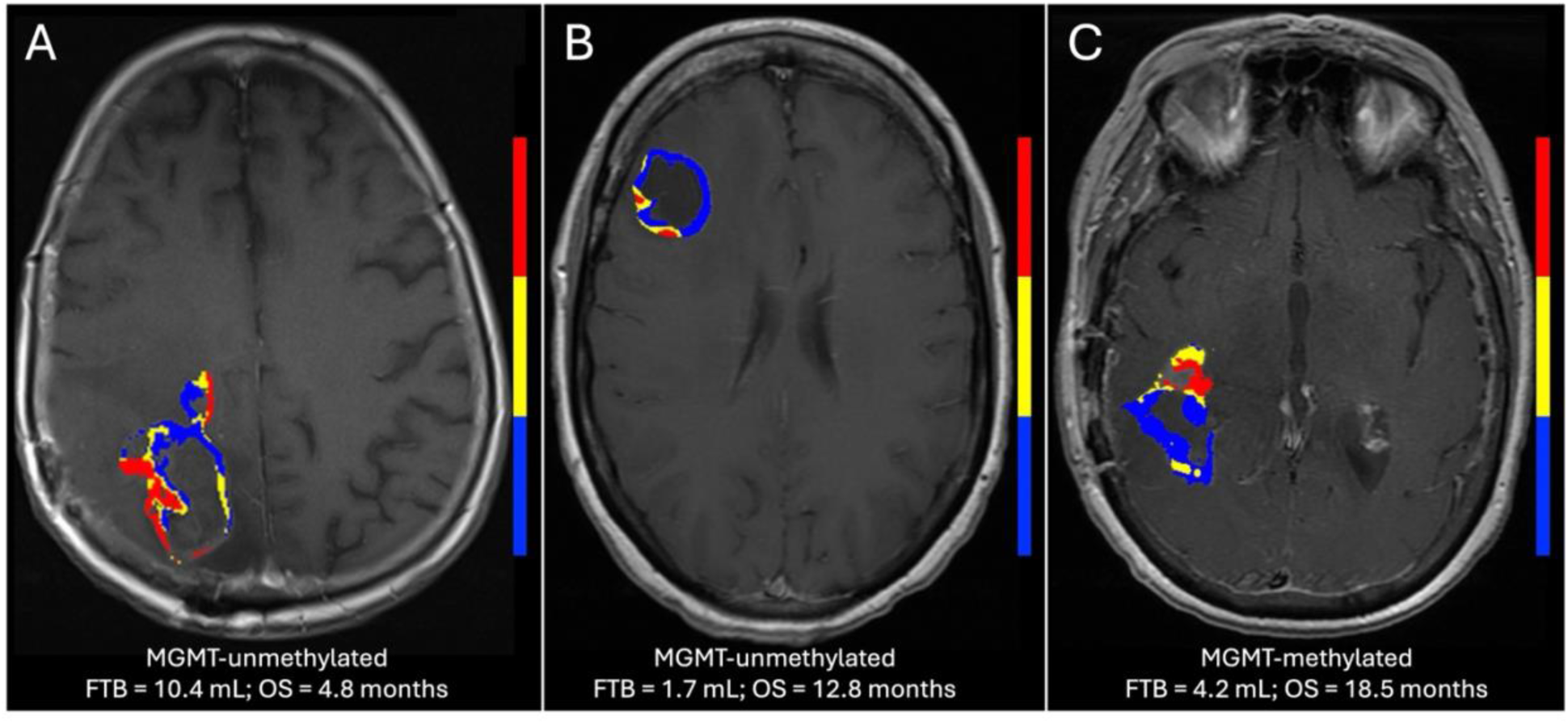
Representative examples of fractional tumor burden (FTB) maps demonstrating variability in perfusion-defined tumor burden and associated clinical outcomes. A-B, MGMT-unmethylated tumors showing (A) higher FTB (red and yellow regions) corresponding to shorter OS and (B) lower FTB corresponding to longer OS. C, Example of an MGMT-methylated tumor demonstrating longer OS with relatively low FTB.

## DISCUSSION

In this study, fractional tumor burden (FTB) provided prognostic stratification beyond MGMT methylation status in patients with glioblastoma within six weeks of chemoradiotherapy completion. Among MGMT-unmethylated tumors, patients with low FTB demonstrated significantly longer OS compared with those with high FTB (median OS, 14.46 vs 8.39 months; P = .006), with OS approaching that of MGMT-methylated tumors. In the present cohort, median OS was 19.67 months for MGMT-methylated tumors and 10.59 months for MGMT-unmethylated tumors, reflecting a separation similar to previously published research.^1^ Given that MGMT methylation is one of the most established prognostic markers in glioblastoma, these findings indicate that perfusion-defined tumor burden captures biologic heterogeneity within the MGMT-unmethylated population and may help explain variability in outcomes within this traditionally poor-prognosis group.

The clinical relevance of the low-FTB MGMT-unmethylated subgroup should be interpreted in the context of MGMT status as an important but incomplete predictor of treatment response. MGMT-unmethylated glioblastomas are typically associated with reduced benefit from temozolomide; however, this classification does not imply uniform treatment resistance across all patients. Prior studies have reported outcomes suggesting that some patients with MGMT-unmethylated tumors may still benefit from temozolomide-based therapy.^9^ In addition, prior work has shown that perfusion-defined tumor vascularity provides prognostic information complementary to MGMT methylation status.^34^ Prior work has suggested that the relationship between perfusion metrics and treatment response is complex, with elevated pretreatment perfusion associated with treatment response in some cohorts.^35^ In this context, the ability of FTB to stratify survival within the MGMT-unmethylated population suggests that spatially resolved perfusion-defined tumor burden may capture clinically relevant variation not fully reflected by binary MGMT classification, including the possibility of low-level MGMT promoter methylation or other favorable treatment-related biology.

These results support the concept that contrast enhancement on postcontrast T1WI following chemoradiotherapy is not biologically uniform, particularly within the early post-treatment period. While conventional postcontrast T1WI reflects disruption of the BBB, it does not reliably distinguish between viable vascular high-grade tumor and treatment-related changes, especially within the first few months following treatment when pseudoprogression is most commonly observed. In contrast, perfusion imaging enables characterization of enhancement based on underlying vascularity, with elevated perfusion associated with viable tumor burden. In this context, fractional tumor burden provides a spatial measure of the biologically active portion of the enhancing region, offering insight beyond conventional imaging.

Advanced imaging metrics, including sRCBV and ADC, were not associated with OS in this cohort, which may reflect the use of whole-lesion regions of interest that do not capture underlying spatial heterogeneity. Similarly, changes in enhancement on postcontrast T1WI following chemoradiotherapy were not associated with OS, likely because enhancement reflects a range of underlying processes related to BBB disruption rather than viable tumor alone.

The focus on early post-treatment imaging represents an additional clinically relevant aspect of this study. Imaging was performed within six weeks following chemoradiotherapy, a period during which interpretation of contrast enhancement is particularly challenging due to pseudoprogression, which may occur within the first 3-4 months after treatment completion. As a result, many clinical trials require extended follow-up to confirm progression, often delaying response assessment and enrollment. In this context, imaging approaches that better characterize early post-treatment enhancement may provide value in distinguishing viable tumor from treatment-related change and could help inform clinical decision-making during this critical interval.

This study has several limitations. It is a retrospective, single-institution analysis with a relatively small sample size, which may limit generalizability. In addition, imaging-defined regions were not correlated with histopathologic sampling, and therefore the biologic interpretation of perfusion-defined tumor burden remains inferential. Multivariable analysis was not performed given the limited sample size and covariate distribution, and the findings should be interpreted in the context of unadjusted associations. Furthermore, ADC was evaluated using whole-lesion regions of interest, which may mask underlying spatial heterogeneity, and more advanced diffusion techniques were not assessed. Finally, while tumor vascularity has been associated with tumor grade, aggressiveness, and treatment response in multiple settings, this analysis focused on the early post-chemoradiotherapy period. As such, the interpretation and prognostic significance of FTB may differ in other treatment contexts. Further studies in larger, prospective cohorts are needed to validate these findings and further define the role of perfusion-based imaging in post-treatment assessment.

## CONCLUSIONS

Fractional tumor burden provides prognostic stratification beyond MGMT methylation status in glioblastoma, identifying a subgroup of MGMT-unmethylated tumors with improved OS following standard chemoradiotherapy. These findings support that post-treatment contrast enhancement is biologically heterogeneous and can be further characterized using spatially resolved perfusion-based imaging. This approach may improve interpretation of early post-treatment imaging by distinguishing viable tumor from non-tumor-related enhancement.

Together, these findings suggest that FTB may help identify clinically meaningful outcome differences among patients with MGMT-unmethylated glioblastoma following temozolomide-based chemoradiotherapy.

## Data Availability

Deidentified data supporting the findings of this study are available from the corresponding author upon reasonable request, subject to institutional approval and applicable data-sharing restrictions.

## Author Contributions

Substantial contributions to all aspects of this work, including conception, analysis, interpretation, writing, and final approval (MAP, JMC, KMS).

## Previously presented in part

- Prah, MA, Connelly, JM, and Schmainda, KM. MGMT combined with post-CRT perfusion MRI-derived Fractional Tumor Burden informs overall survival in newly diagnosed glioblastoma. Annual Meeting of the American Society of Neuroradiology (ASNR) New York City, NY, USA. 2022. **2022 Adult Brain Outstanding Presentation Award.**
- Prah, MA, Connelly, JM, and Schmainda, KM. Unmethylated MGMT combined with post-CRT perfusion MRI-derived tumor burden informs overall survival in newly diagnosed glioblastoma. 30th Annual Meeting of the International Society of Magnetic Resonance in Medicine (ISMRM), London, England, UK. 2022.
- Prah, MA, Connelly, JM, and Schmainda, KM. Low Fractional Tumor Burden volume obtained from perfusion MRI reveals significant survival benefit in MGMT unmethylated glioblastoma. Annual Meeting of the Society for Neuro-Oncology (SNO). Tampa, Florida, USA. 2022.

## ABBREVIATIONS

CRT: concomitant chemoradiation therapy
dT1: delta T1
FTB: fractional tumor burden
IDH: isocitrate dehydrogenase
MGMT: O^6^-methylguanine DNA methyltransferase
OS: overall survival
sRCBV: standardized relative cerebral blood volume

